# Anti-spike protein receptor-binding domain IgG levels after COVID-19 infection or vaccination against SARS-CoV-2 in a seroprevalence study

**DOI:** 10.1101/2021.06.06.21258406

**Authors:** Hiie Soeorg, Piia Jõgi, Paul Naaber, Aigar Ottas, Karolin Toompere, Irja Lutsar

## Abstract

**Purpose:** In a country-wide seroprevalence study of COVID-19 in Estonia we aimed to determine the seroprevalence and the dynamics of IgG against SARS-CoV-2 after vaccination or positive PCR-test.

**Methods:** Leftover blood samples were selected between February 8 to March 25, 2021, by SYNLAB Estonia from all counties and age groups (0-9, 10-19, 20-59, 60-69, 70-79, 80-100 years) proportionally to the whole Estonian population and tested for IgG against SARS-CoV-2 spike protein receptor-binding domain (anti-S-RBD IgG) using Abbott SARS-CoV-2 IgG II Quant assay. Antibody levels after positive PCR-test or vaccination were described by nonlinear model.

**Results:** A total of 2517 samples were tested. Overall seroprevalence (95% CI) was 20.1% (18.5-21.7%), similar in all age groups. If all individuals vaccinated with the first dose at least 14 days before antibody measurement were assumed to be seronegative, the overall seroprevalence was 15.8% (14.4-17.3%), 4-fold larger than the proportion of confirmed COVID-19 cases. According to nonlinear models, age increased anti-S-RBD IgG production after positive PCR-test but decreased after vaccination. The peak of anti-S-RBD IgG in a 52-year-old (median age of PCR-positive and/or vaccinated individuals) was significantly higher after vaccination compared with positive PCR-test (22082 (12897…26875) vs 6732 (2321…8243) AU/mL), but half-life was similar (26.5 (6.9…46.1) vs 38.3 (8.2…68.5) days).

**Conclusion:** One year after the start of COVID-19 pandemic the actual prevalence of infection is still underestimated compared with confirmed COVID-19 cases, underlining the importance of seroepidemiological studies. Older individuals have lower anti-S-RBD IgG level after vaccination, but similar decline rate to younger.

## Introduction

On December 27, 2020, Estonia started a national vaccination program against COVID-19 with Pfizer/BioNTech vaccine, followed by Moderna and AstraZeneca vaccine in January and February 2021, respectively. Initially, the vaccines were administered mostly to healthcare workers and older people (at least 80 years old). By February 7, 2021, 1.4% of the whole population had been vaccinated with one and 1.3% with two doses [1]. In clinical trials, the immunological response to vaccines has been well described [2, 3], but real-world data about IgG levels is limited. However, determining the variables affecting the antibody response, particularly older age as one of the most important risk factors for severe COVID-19 [4], in a real-world setting is relevant to answer questions that are not addressed in the trials to improve age-targeted strategies to prevent the disease.

Compared with seroprevalence of 1.5% or 6.8% in two different counties between May to July 2020 [5], by February 7, 2021, the overall cumulative proportion of COVID-19 cases confirmed by SARS-CoV-2 PCR in Estonia was 3.5% [1]. As the ratio of the actual prevalence of infection to confirmed cases of COVID-19 can vary widely up to 23 times [5, 6], seroepidemiological studies are warranted to determine the seroprevalence and thereby individuals already immune to the disease. Therefore, a country-wide cross-sectional seroepidemiological study of COVID-19, KoroSero-EST-3, was conducted to determine the proportion of individuals seropositive to SARS-CoV-2 in Estonia.

In this study, we aimed to determine the seroprevalence and the dynamics of IgG antibodies against SARS-CoV-2 after vaccination or positive PCR-test in Estonia one year after the beginning of the pandemic. Additionally, we assessed whether the IgG levels differ between different age groups.

## Methods

### Study design

The KoroSero-EST-3 was a cross-sectional seroepidemiological study conducted from February 8 to March 25, 2021, in Estonia. Leftover blood samples were selected by SYNLAB Estonia from samples sent by general practitioners for routine clinical testing from all fifteen counties and age groups (0-9, 10-19, 20-59, 60-69, 70-79, 80-100 years) proportionally to the whole Estonian population. Data on positive PCR-tests and SARS-COV-2 vaccination history were obtained from The Electronic Health Record, a nationwide system collecting data from healthcare providers in Estonia that each patient can access online, by Health and Welfare Information Systems Centre. The study was approved by the Research Ethics Committee of the University of Tartu. Informed consent was waived.

### Antibody measurement

Blood samples containing at least 500 μL after routine clinical tests were selected according to the sample size calculations. All samples were tested for IgG antibodies against SARS-CoV-2 spike protein receptor-binding domain (anti-S-RBD IgG) by chemiluminescent microparticle immunoassay (Abbott SARS-CoV-2 IgG II Quant with ARCHITECT i2000SR analyzer; Abbott Laboratories, USA) according to the manufacturer’s instructions. A sample was considered positive for anti-S-RBD IgG if the result was ≥50 AU/mL.

### Statistical analysis

The software program Stata 14.2 was used for calculating sample size and the total number of individuals with IgG antibodies and R (version 3.6.3) for other statistical analyses. Sample size per age group in each county was calculated to obtain binomial exact 95% confidence interval (CI) with the width of 10% for seroprevalence estimate in each age group, assuming seroprevalence was 10%. A total of 2609 samples was needed.

Seroprevalence was estimated as the proportion of samples positive for anti-S-RBD IgG, overall and in each county and age group separately. Binomial exact 95% CI was calculated for seroprevalences. The total number of individuals with IgG antibodies was estimated by taking into account stratification to age groups and counties.

An individual was considered as seropositive due to a positive PCR-test or vaccination with the first dose if PCR-test or vaccination with the first dose occurred at least 14 days prior to antibody measurement. Multinomial 95% CI was calculated for the proportion of seropositive individuals with positive PCR-test and/or vaccinated with the first or second dose. Antibody levels between PCR-positive and/or vaccinated individuals were compared with pairwise Wilcoxon tests with Holm adjustment for multiple comparison.

Antibody levels *C*(*t*) at time *t* after positive PCR-test or the first dose of Pfizer/BioNTech or Moderna vaccine was described by exponential increase (with production rate *a*) to peak level of 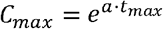 at time *t*_*max*_ followed by exponential decline (with decline rate *b*), fitted by non-linear least squares method:

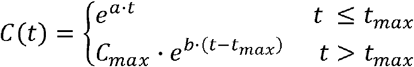

The effect of age on parameters *a, b, t*_*max*_ was tested by multiplying the parameter by 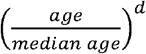 or (1 + (*age* – *median age*)· *d*. Covariance matrix of parameters was calculated by applying Eicker-White method to handle heteroscedasticity using R package regtools [7]. Monte Carlo simulation was used to calculate 95% CI for peak level *C*_*max*_ and mean antibody level using R package propagate [8]. Antibody half-life in days was calculated as 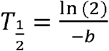.

Daily numbers of COVID-19 cases confirmed by PCR-test for SARS-CoV-2 from nasopharyngeal swab, deaths due to COVID-19 and individuals vaccinated in each county and age group were obtained from Estonian Health Board [1] and the number of inhabitants by county and age group as of January 1, 2021, from Statistics Estonia [9]. The total cumulative number of confirmed COVID-19 cases and individuals vaccinated with two doses per 100 inhabitants was calculated as of two weeks before the midpoint (March 2, 2021) of sample collection (February 16, 2021). To compare the cumulative proportion of confirmed COVID-19 cases with seroprevalence, seroprevalence excluding vaccinated, i.e. assuming individuals vaccinated with the first dose at least 14 days prior to antibody measurement were seronegative, was calculated. Infection fatality rate was calculated as number of deaths due to COVID-19 as of February 16, 2021, divided by the estimated number of seropositive individuals excluding vaccinated. To calculate confidence intervals of infection fatality rate the number of deaths was assumed to follow Poisson distribution.

## Results

A total of 2517 samples from individuals with distribution into age groups and counties of residence similar to the whole Estonian population were analyzed (Table 1). During the sample collection period, the proportion of individuals who had had positive PCR-test or had been vaccinated with two doses rose from 3.6% to 7.5% and 1.4% to 4.5%, respectively (Fig. 1).

**Table 1.** Age and county of residence of the study population and the whole Estonian population, confirmed COVID-19 cases, and seroprevalence with 95% confidence interval (CI)

|  | Study population (n=2517) | The whole population (%) | Confirmed COVID-19 cases in the whole population (%) | Seroprevalence (%) (95% CI) | Seroprevalence excluding vaccinated (%) (95% CI) | Ratio of seropositive excluding vaccinated to confirmed COVID-19 cases |
| --- | --- | --- | --- | --- | --- | --- |
| **Age group** |  |  |  |  |  |  |
| 0-9 | 205 (8.1%) | 10.8 | 1.2 | 19.5 (14.3-25.6) | 19.5 (14.3-25.6) | 16.3 |
| 10-19 | 262 (10.4%) | 10.4 | 4.0 | 19.1 (14.5-24.4) | 19.1 (14.5-24.4) | 4.8 |
| 20-59 | 1333 (53.0%) | 52.3 | 4.8 | 20.2 (18.1-22.4) | 16.7 (14.7-18.8) | 3.5 |
| 60-69 | 336 (13.3%) | 12.3 | 4.2 | 19.3 (15.3-24.0) | 15.5 (11.8-19.8) | 3.7 |
| 70-79 | 225 (8.9%) | 8.4 | 2.9 | 17.8 (13.0-23.4) | 11.1 (7.32-16.0) | 3.8 |
| $\geq 80$ | 156 (6.2%) | 5.8 | 4.7 | 26.9 (20.1-34.6) | 5.1 (2.2-9.9) | 1.1 |
| **County** |  |  |  |  |  |  |
| Harju | 1158 (46.0%) | 45.8 | 4.7 | 28.0 (25.4-30.7) | 22.4 (20-24.9) | 4.8 |
| Hiiu | 20 (0.8%) | 0.7 | 3.3 | 10.0 (1.2-31.7) | 10.0 (1.2-31.7) | 3.0 |
| Ida-Viru | 254 (10.1%) | 9.9 | 7.2 | 17.7 (13.2-23.0) | 16.5 (12.2-21.7) | 2.3 |
| Jõgeva | 53 (2.1%) | 2.1 | 2.2 | 13.2 (5.5-25.3) | 9.4 (3.1-20.7) | 4.3 |
| Järva | 59 (2.3%) | 2.2 | 2.5 | 5.1 (1.1-14.2) | 5.1 (1.1-14.1) | 2.0 |
| Lääne | 42 (1.7%) | 1.5 | 1.6 | 14.3 (5.4-28.4) | 0 (0-8.4) | 0.0 |
| Lääne-Viru | 108 (4.3%) | 4.4 | 2.2 | 4.6 (1.5-10.5) | 3.7 (1.0-9.2) | 1.7 |
| Põlva | 50 (2.0%) | 1.8 | 1.8 | 16.0 (7.2-29.1) | 10.0 (3.3-21.8) | 5.6 |
| Pärnu | 148 (5.9%) | 6.4 | 3.0 | 13.5 (8.5-20.1) | 7.4 (3.8-12.9) | 2.5 |
| Rapla | 65 (2.6%) | 2.5 | 3.0 | 9.2 (3.5-19.0) | 7.7 (2.5-17) | 2.6 |
| Saare | 62 (2.5%) | 2.5 | 3.6 | 12.9 (5.7-23.9) | 9.7 (3.6-19.9) | 2.7 |
| Tartu | 285 (11.3%) | 11.6 | 2.5 | 16.5 (12.4-21.3) | 12.6 (9.0-17.1) | 5.0 |
| Valga | 57 (2.3%) | 2.1 | 2.4 | 10.5 (4.0-21.5) | 7.0 (1.9-17.0) | 2.9 |
| Viljandi | 85 (3.4%) | 3.4 | 1.7 | 11.8 (5.8-20.6) | 9.4 (4.2-17.7) | 5.5 |
| Võru | 71 (2.8%) | 2.6 | 3.3 | 12.7 (6.0-22.7) | 9.9 (4.1-19.3) | 3.0 |

**Fig. 1.**
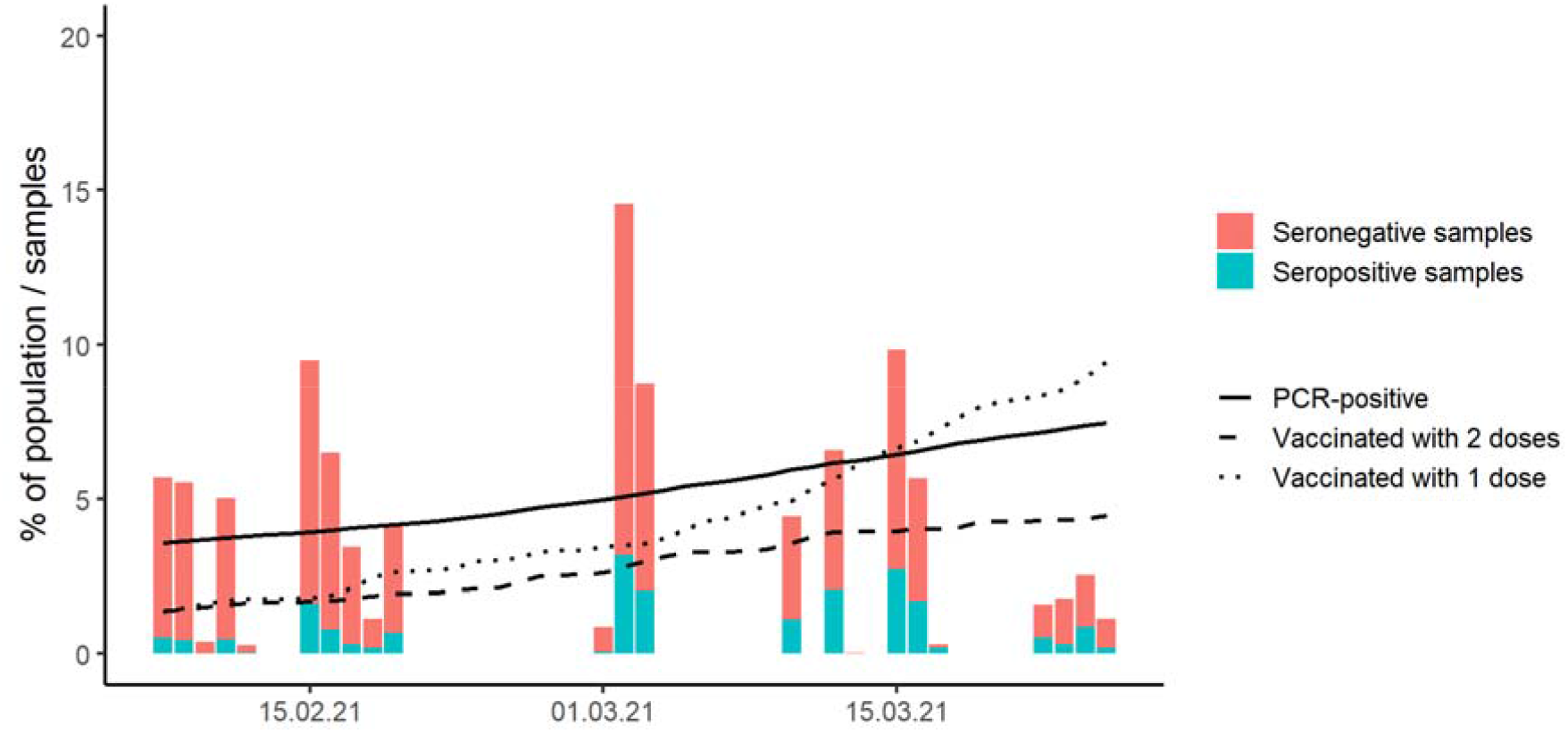
Timing of collection of leftover blood samples (red – seronegative samples, green – seropositive samples), the cumulative proportion of confirmed COVID-19 cases (solid line), vaccinated with two doses (dashed line), and vaccinated with one dose (dotted line) in the whole Estonian population

A total of 207 (8.2%) individuals had positive PCR-test and 48 (1.9%) had been vaccinated with one dose at least 14 days before antibody measurement (10 with AstraZeneca, 34 Pfizer/BioNTech, 4 Moderna), 71 (2.8%) had received two doses (none with AstraZeneca, 68 Pfizer/BioNTech, 3 Moderna) and 3 (0.1%) had been PCR-positive and vaccinated with at least one dose. Of the whole Estonian population, two weeks prior to the midpoint of sample collection 4.0 % had been PCR-positive, 1.9% had been vaccinated with one dose, 1.7% vaccinated with two doses.

A total of 506 samples were seropositive; overall seroprevalence was (95% CI) was 20.1% (18.5-21.7%) and the estimated number of seropositive individuals in Estonia 265 585 (95% CI 245 452-285 718). Seroprevalence was similar in all age groups but varied between counties, being significantly larger in some counties, particularly Harju county where the capital Tallinn is located and that was the focus region of second wave of infections (Table 1).

If all individuals vaccinated with the first dose at least 14 days prior to antibody measurement were assumed to be seronegative, the overall seroprevalence was 15.8% (95% CI 14.4-17.3%), smallest in individuals ≥80 years old (Table 1). The ratio of seropositive individuals excluding vaccinated to confirmed COVID-19 cases was 4.0, ranging in various age groups up to 16.3, being largest in children <10 years old, and in counties up to 5.6. A total of 508 individuals had died due to COVID-19 as of February 2021, yielding infection fatality rate 0.24% (95% CI 0.21-0.27%).

Overall, of seropositive individuals 200 (39.5%; 95% CI 35.0-44.3%) had had positive PCR-test and 44 (8.7%; 4.2-13.4%) vaccinated with one dose at least 14 days prior to antibody measurement, 65 (12.8%; 8.3-17.6%) with two doses, 3 (0.6%; 0.0-5.3%) had been PCR-positive and vaccinated with at least one dose, 194 (38.3%; 33.8-43.1%) had not had positive PCR-test or been vaccinated. These proportions varied between age groups – seropositive children aged 0-9 years mostly had not been PCR-positive, seropositive adolescents and adults up to 79 years mostly had been PCR-positive, at least 80 years old seropositive individuals had mostly been vaccinated (Fig. 2).

**Fig. 2.**
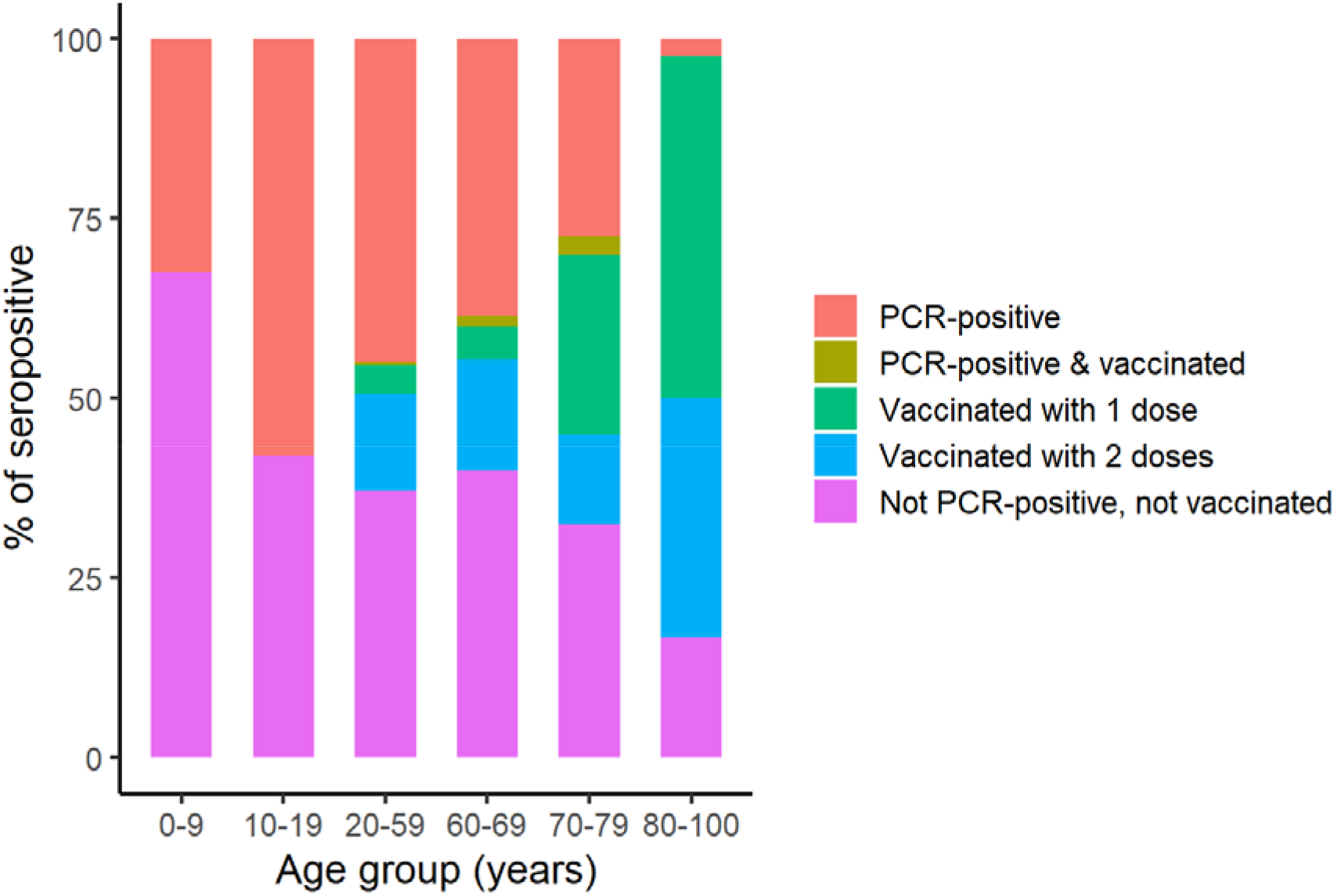
The proportion of PCR-positive, PCR-positive and vaccinated with at least one dose, vaccinated with one or two doses, not PCR-positive and not vaccinated individuals among seropositive individuals by age group

Seropositive individuals comprised 96.6% (95% CI 93.2-9.6%) of those with positive PCR-test and 91.7% (80.0-97.7%) vaccinated with one dose at least 14 days prior to antibody measurement, 91.5% (82.5-96.8%) vaccinated with two doses, 100% (29.2-100%) with positive PCR-test and vaccinated with at least one dose, 8.9 (7.7-10.1%) others. Median (IQR) time between antibody measurement and positive PCR-test was 37 (22-73) days, vaccination with one dose 21 (19-21) and with two doses 14 (5.5-27) days. Anti-S-RBD levels in those vaccinated with two doses (median (IQR) 7682 (10469-23400) AU/mL) was larger than in PCR-positive individuals (1039 (390-2633) AU/mL) (p=0.022), those vaccinated with one dose (289 (89-781) AU/mL) (p<0.001), not PCR-positive and not vaccinated individuals (528 (154-1903) AU/mL) (p=0.005), but not in PCR-positive and vaccinated with at least one dose (13732 (10469-23400) AU/mL) (p=0.223) (Fig. 3).

**Fig. 3.**
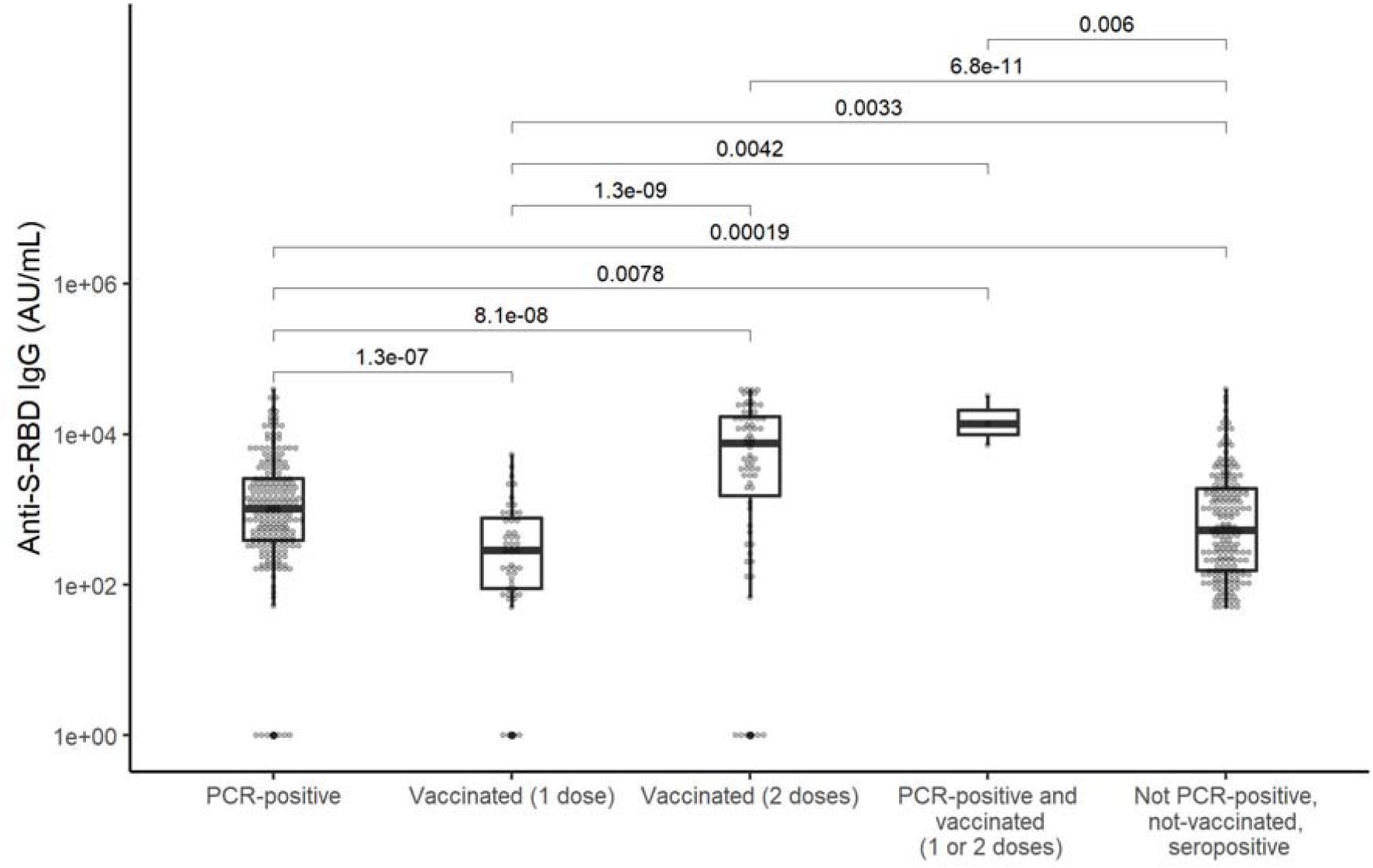
IgG levels against receptor binding domain of spike protein (anti-S-RBD IgG) in PCR-positive and/or vaccinated individuals and not PCR-positive, not vaccinated, but seropositive individuals. The numbers above the boxplots show statistically significant p-values (<0.05) from pairwise Wilcoxon tests with Holm adjustment for multiple comparison

A non-linear model of anti-S-RBD IgG levels was developed separately for all individuals with positive PCR-test (n=278) and vaccinated with one dose of Pfizer/BioNTech or Moderna vaccine (n=56 and n=7, respectively) or two doses (n=68 and n=3, respectively). The time between one and two doses of Pfizer/BioNTech or Moderna vaccine was median (IQR) of 21 (21-22) and 28 (28-28) days, respectively. The peak level of antibodies occurred 17 days after positive PCR-test or 32 days after the first dose of vaccine (Table 2). Increasing age increased antibody production after positive PCR-test, but decreased after vaccination as shown by parameter *d* (Table 2, Fig. 4). Modelled peak level in a 52-year-old (median age of PCR-positive and/or vaccinated individuals) individual was significantly higher after vaccination compared with positive PCR-test, but antibody decline rate *b* and half-life were similar (Table 2).

**Table 2.** Parameters and 95% confidence interval (CI) of non-linear models describing production and decline of IgG levels against receptor binding domain of spike protein (anti-S-RBD IgG) after positive PCR-test or vaccination with Pfizer/BioNTech or Moderna vaccine

| Parameter | Model for anti-S-RBD IgG levels |  |
| --- | --- | --- |
|  | after positive PCR-test<br>Estimate (95% CI) | after vaccination<br>Estimate (95% CI) |
| Antibody production rate: |  |  |
| $a \cdot (1 + (age - 52) \cdot d)$ | | |
| $a$ (1/day) | 0.51 (0.45...0.57) | 0.31 (0.30...0.33) |
| $d$ | 0.004 (0.001...0.006) | -0.001 (-0.001...-0.0004) |
| Antibody decline rate $b$ (1/day) | -0.019 (-0.31...-0.007) | -0.026 (-0.046...-0.007) |
| Time when peak level occurs $t_{max}$ (days) | 17.2 (15.1...19.4) | 32.0 (30.2...33.8) |
| Peak level at time $t_{max}$ at age 52 years (AU/mL) | 6732 (2321...8243) | 22082 (12897...26875) |
| Half-life (days) | 38.3 (8.2...68.5) | 26.5 (6.9...46.1) |

**Fig. 4.**
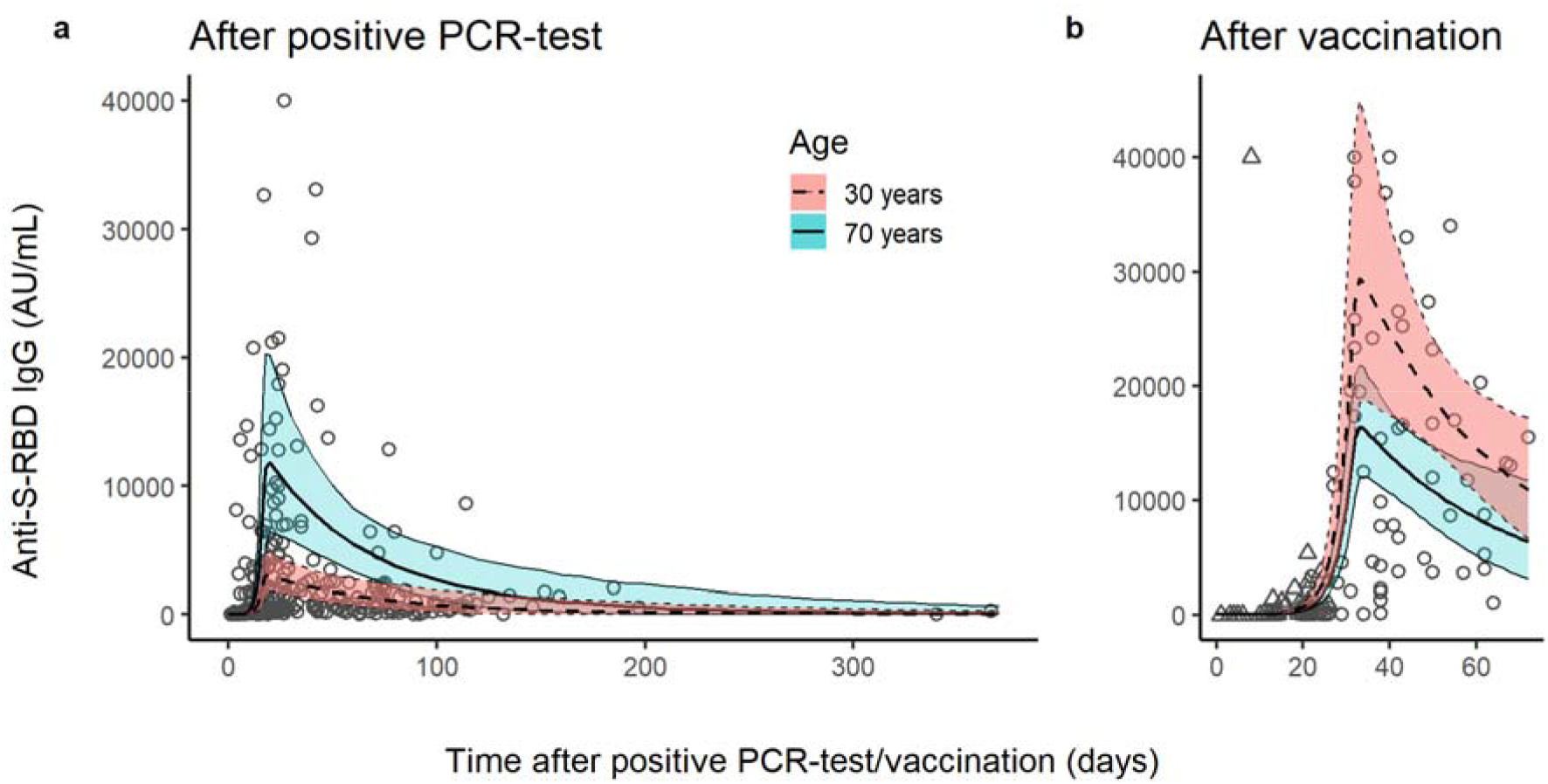
Mean IgG level against receptor binding domain of spike protein (anti-S-RBD IgG) and 95% confidence interval in a 30-year-old individual (dashed lines, red shaded area) or a 70-year-old individual (solid lines, green shaded area) after a) positive PCR-test or b) vaccination with the first dose. Dots show observed antibody levels in those with positive PCR-test or vaccinated with two doses, triangles in those vaccinated with one dose

## Discussion

To the best of our knowledge, this is the first country-wide seroepidemiological study describing in addition to the seroprevalence the dynamics of anti-S-RBD IgG after positive PCR-test or vaccination. Seroprevalence of COVID-19 one year after the start of the pandemic and about 2 months after the start of vaccination was 20.1%, similar in all age groups. After positive PCR-test older individuals had higher peak level of anti-S-RBD IgG and after vaccination lower levels compared with younger individuals. The decline rate of antibodies was similar regardless of age.

The seroprevalence of SARS-CoV-2, 20.1% in February/March 2021, is considerably higher compared to 1.5-6.3% in May to July 2020 [5], as expected. Similar seroprevalence, 19%, was found in Sweden in February 2021 [10] where the cumulative proportion of confirmed COVID-19 cases was 6.5% [11], somewhat higher than in Estonia (4.0%). Several other results of our seroepidemiological study are also consistent with the spread and nature of the virus and the strategies to prevent it. First, the ratio of seroprevalence excluding vaccinated to the proportion of confirmed COVID-19 cases ranged up to 5.6 that is less than up to 13.3 during the first wave of the pandemic in Estonia [5], possibly due to increased PCR-testing rate (average daily number of tests per 100 000 inhabitants 64 up to July 31, 2020, vs 347 in 2021 up to February 7, 2021 [1]). Second, the larger proportion of individuals without positive PCR-test among seropositive younger compared with older people once again corroborates that we may miss a considerable number of infections, up to 93% in children [12], when relying only on testing of symptomatic cases due to higher rate of asymptomatic or mildly symptomatic cases among children compared with adults [13]. Finally, the lowest seroprevalence and its ratio to confirmed COVID-19 cases among older people suggest that protecting the elderly from becoming infected and their careful testing was successful, although we cannot exclude the contribution of higher IgG decline rate in older people found in some studies [14, 15]. However, as 73.6% of COVID-19 cases confirmed by February 7, 2021, in Estonia had occurred since December 1, 2020 [1], i.e. within a timeframe of >6 months when anti-S-RBD IgG should persist at detectable levels [16], we believe that waning immunity may have only minor impact on the seroprevalence estimates.

Similarly to other studies, we found that older individuals have higher anti-S-RBD IgG peak levels after infection compared with younger adults [17, 18]. However, such association with age has not been consistently found, as in other studies higher anti-S-RBD IgG correlates with the presence of fever or severity of symptoms instead [16, 19]. In our previous study we also found that after adjusting for age, time since positive PCR-test and sex, hospitalized patients or those with fever had higher anti-RBD IgG levels than those not hospitalized or without fever [5]. As older people have more severe symptoms [4], the relationship between age and antibody peak level may have been mediated by this association. This is supported in our study by lower anti-S-RBD-IgG levels in seropositive individuals without positive PCR-test and not vaccinated, who possibly did not go to testing due to asymptomatic infection.

Despite that faster decline of antibodies against SARS-CoV-2 in older people after the infection has been found [15], the inclusion of age to decline rate of anti-S-RBD IgG did not improve the fit of the models in our study. However, the inverse relationship between age and decline rate has not been reported consistently [16, 19] and similarly to peak level may be instead mediated by the severity of symptoms [14, 16], although this is controversial [15, 19]. Still, in the model of anti-S-RBD IgG after vaccination that was mostly based on data from older people, the half-life was somewhat shorter, but not significantly, and it could be also due to the short observation period of vaccinated individuals as initially more rapid decline occurs compared with later time period [14, 20, 21]. Nevertheless, as after vaccination older people have lower anti-S-RBD IgG peak level, also found by others [22-24], their antibody levels will be lower for some time compared with younger individuals until equalizing [24]. Due to limited data so far, further studies are warranted to determine whether the older age groups, at the highest risk of severe disease, are also at the risk of faster decline of IgG after vaccination.

Several limitations in our study should be noted. First, the study design, use of leftover blood samples, may have led to somewhat biased results due to the risk of including disproportionally more sick people. Indeed, we had a somewhat larger proportion of people who had had positive PCR-test (8.2%) compared with the proportion in the whole population (4.0%). However, as the seroprevalence was similar to 21.4% (19.8-23.1%) estimated in the study on the prevalence of the coronavirus in Estonia based on a random statistical sample conducted between March 11 to March 22, 2021, [25], we believe that potential bias is small. Second, we did not have data about important covariates affecting antibody levels, particularly symptoms and sex [16, 19], and thus we were not able to adjust our analyses for these variables. Still, such impact of the study design is offset by its advantages like rapidity, low cost, no additional burden to healthcare workers or patients, avoiding physical contact that is particularly relevant in the case of COVID-19, to gain insight into the proportion of individuals protected from the disease and their antibody levels. Third, we do not know whether some individuals vaccinated had been infected with SARS-CoV-2 prior to vaccination without a positive PCR-test. As one vaccine dose after prior infection results in antibody levels similar to the titer after two doses administered to those not previously infected [26], the estimates from our model may be somewhat biased. However, as most of the vaccinated people were older people and seroprevalence excluding vaccinated was low among them, we believe that the proportion of individuals with undetected COVID-19 infection prior to vaccination was small.

In conclusion, one year after the start of pandemic about one fifth of Estonian population regardless of age had antibodies against SARS-CoV-2 that is four times larger than estimated by national statistics based on PCR-positivity, underlining the importance of seroepidemiological studies to understand the actual prevalence of infection. Older individuals have lower anti-S-RBD IgG peak levels after vaccination but not after natural infection. Whether lower peak levels result in potentially shorter persistence of antibodies, thus immunity and need for booster doses of the vaccine, requires further long-term studies determining the dynamics of antibodies after vaccination.

## Data Availability

The data of this study are available from the corresponding author upon reasonable request.

## Availability of data and material

The data of this study are available from the corresponding author upon reasonable request.

## Code availability

The code used in the analyses of this study is available from the corresponding author upon reasonable request.

## Ethics approval

The study was approved by the Research Ethics Committee of the University of Tartu.

## Consent to participate

Informed consent was waived.

## Consent for publication

Informed consent was waived.

## Acknowledgements

We thank Health and Welfare Information Systems Centre and all the study participants. Electronic data collection was supported by the High Performance Computing Center of the University of Tartu.

